# Pilot study of a high-intensity interval training program in older adults: Safety, feasibility, functional fitness and cognitive effects

**DOI:** 10.1101/2024.01.09.23299774

**Authors:** Vanessa R. Anderson, Katherine Kakuske, Christian Thompson, Maria V. Ivanova

## Abstract

Exercise can boost physical and cognitive health in older adults. However, there are a lack of accessible exercise programs that foster adherence among older adults. In this study, we aimed to establish the safety and feasibility of APEX, a new exercise program designed to optimize fitness and cognitive gains for older adults, in addition to evaluating its acute physiological effects, and assessing its possible effects on functional fitness and cognition among healthy older adults. APEX utilizes a multimodal progressive high-intensity interval training (HIIT) design, with high-intensity intervals focused on enhancing cardiovascular fitness and muscle strength, and recovery intervals that incorporate balance and mobility exercises. The APEX training was tested in healthy older adults (n=4) over the course of four weeks. Ultimately, APEX was found to be safe and feasible, with no adverse events and high adherence. Participants met heart rate targets for all of the high-intensity exercises, and all intervals had a significant difference in heart rates between high-intensity and recovery periods in linear effects models (p<0.001). Improvements in functional fitness were observed in aerobic endurance, lower body strength, and balance. The intervention was also associated with positive trends in the cognitive domains of information processing, working memory, executive control, and attention. APEX offers a promising alternative to traditional cardiovascular exercise modalities for older adults with additional benefits for functional fitness and cognition. These results encourage further testing of the APEX program in older adults and different clinical populations.

## Introduction

The demographic of older adults aged 65 years and older is rapidly growing worldwide, projected to reach 1.6 billion in 2050 (World Population Ageing 2019, 2020). Aging is accompanied by declines in physical health and cognitive functioning (Physical Activity Guidelines Midcourse Report, 2023). These age-related declines lower the quality of life for older adults and burden the healthcare system and the economy (Physical Activity Guidelines Midcourse Report, 2023). Physical activity is an accessible and affordable intervention that has been used to boost physical health and cognition in older adults (Langhammer et al., 2019; Physical Activity Guidelines Midcourse Report, 2023). Some physical benefits of exercise in older adults include improved muscle strength and power, balance, gait speed, and stride length (Gibala et al., 2018; Marriott et al., 2021). The observed cognitive benefits of exercise include improvements in executive function and working memory (Klimova & Dostalova, 2020; Xiong et al., 2021; Zhidong, 2021). Additionally, regular exercise protects against the onset of or progression of age-related diseases, such as cardiovascular and neurodegenerative disorders (Duzel et al., 2019; Fuller et al., 2020).

The Centers for Disease Control and Prevention advises older adults to engage in 150 minutes per week of moderate-intensity aerobic exercise or 75 minutes per week of vigorous-intensity aerobic exercise, in addition to two days per week of strength training and multicomponent exercises that target balance and flexibility (Centers for Disease Control and Prevention, 2023). Adhering to these guidelines protects older adults’ long-term health by reducing the risk of chronic diseases and their progression, while also improving functional independence (Elsawy & Higgins, 2010). Alarmingly, though, less than 15% of older adults meet these guidelines (Physical Activity Guidelines Midcourse Report, 2023). The ability of older adults to meet these guidelines is often constrained by age-related functional declines and social and cognitive limitations (Rivera-Torres et al., 2019). Given the rapidly expanding older adult population and low rates of exercise adherence for this population, there is a critical need for physical activity programs that are effective and accessible for older adults. The current study proposes a novel exercise intervention—the APEX program—specifically designed to address existing barriers to participation and enhance adherence for older adults and clinical populations. In this paper, we detail the APEX program and evaluate its feasibility in a cohort of healthy older adults.

In recent studies, High-Intensity Interval Training (HIIT) has emerged as a compelling exercise format with documented physical and cognitive benefits for older adults (Gibala et al., 2018; Marriott et al., 2021; Mekari et al., 2020). HIIT is characterized by repeated intervals of vigorous aerobic exercise followed by short recovery periods of low-intensity exercise (Snarr, 2022). For older adults, HIIT outshines typical Moderate-Intensity Continuous Training (MICT) by offering a multitude of diverse benefits. MICT refers to consistent and sustained exercise at 55-65% Max HR, typically for 30 to 50 minutes, depending on the physical fitness of the participant (Bullock et al., 2020, Coswig et al., 2020). In comparison to MICT, HIIT promotes significantly greater improvements in functional movement in both healthy and clinical older adults (Snarr, 2022; Stern et al., 2023). HIIT has also shown notable enhancements in the cognitive domains of executive functioning and working memory in cognitively intact healthy older adults that surpass the outcomes observed with MICT (Kovacevic et al., 2020; Mekari et al., 2020). The potential benefits of HIIT have been proposed to extend to clinical populations as well, such as stroke patients (Hugues et al., 2021; Kovacevic et al., 2021; Mekari et al., 2020).

The success of HIIT for older adults has been attributed to its high adherence rates, driven by its superior time efficiency relative to MICT (Atakan et al., 2021; Marriott et al., 2021). Additionally, HIIT has been demonstrated to be equally safe and more enjoyable than MICT (Elboim-Gabyzon et al., 2021; Marriott et al., 2021). This assertion gains additional support from the comparable tolerance of HIIT and MICT among older adults with neurodegenerative disorders, as indicated by adverse events, dropouts, and compliance rates (Marriott et al., 2021).

In sum, HIIT is a safe and enjoyable exercise format that offers diverse physical and cognitive benefits to a variety of older adult populations. However, despite these advantages, there is a paucity of accessible HIIT regimens tailored for older adults, and existing regimens do not directly target important fitness domains, like balance and mobility, that are crucial for sustaining physical well-being in older adults (Stern et al., 2023; Marriott et al., 2021).

The novel APEX program introduced in this study leverages the HIIT format to provide similar or even greater physical and cognitive benefits as traditional continuous aerobic exercise regimens, while still being safe and feasible for individuals who may have limited mobility, such as older adults and various clinical populations. Furthermore, APEX is well-suited for group settings and eliminates the need for specialized equipment, thereby increasing its accessibility and affordability. Together, these features foster adherence.

Although most HIIT programs focus on aerobic exercises such as running or cycling, those that incorporate strength, balance, and resistance exercises yield broader physical health benefits extending beyond cardiorespiratory fitness, which is especially appealing to the fitness needs of older adults (Gaedtke & Morat 2015; García-Pinillos et al., 2019; Gibala et al., 2018; Stern et al., 2023). Research on non-HIIT exercises employing multimodal progressive approaches has demonstrated improvements in strength, balance, and gait speed among healthy older adults (Gaedtke & Morat, 2015; Thompson et al., 2019; Morat et al., 2021). These findings strongly suggest that integrating multimodal progressive exercises into a HIIT protocol can complement the inherent advantages of HIIT, maximizing the protocol’s benefits for older adults. In the APEX program, high-intensity intervals emphasize strength training, while the recovery periods uniquely integrate neuromotor (e.g., balance, coordination, sensory stimulation) exercises to optimize cognitive and physical benefits for older adults.

Intensity targets for HIIT programs are based on maximum oxygen consumption (VO_2_Max) and/or maximum heart rate (MaxHR). During a high-intensity interval, the aim is to achieve a VO_2_ between 90% and 120% of VO_2_Max and/or a heart rate (HR) above 80% to 90% of MaxHR. Conversely, recovery interval VO_2_ should ideally fall within the range of 45% to 60% of VO_2_Max, and/or HR should be around 70% of MaxHR (Marriott et al., 2021; Snarr, 2022). It is important to note that the decoupling of VO_2_ and HR responses may occur if major muscle groups are not consistently engaged during high-intensity intervals. This occurrence is observed in some high-intensity resistance training programs that meet their HR targets but not their VO_2_ targets (Ratamess et al., 2015; Snarr, 2022). This is thought to be due to suboptimal oxygen consumption in large muscle groups and increased levels of epinephrine and norepinephrine associated with resistance training, which causes HR to increase faster than VO_2_. To address this concern, APEX incorporates repetitive strength and endurance exercises targeting large muscle groups in its high-intensity periods.

Another parameter of HIIT interventions is the Work-to-Rest Ratio (WRR), the ratio of time engaged in high-intensity exercise to that in recovery (Seo et al., 2019). WRR in HIIT protocols vary widely, depending on the physical traits of participants (e.g. age, functional fitness, health status) (Alzar-Teruel et al., 2022; Marriott et al., 2021; Taylor, 2019). HIIT sessions among older adults typically range from 1:3 to 1:1 WRR (Elboim-Gabyzon et al., 2021; Marriott et al., 2021; Snarr, 2022). The APEX HIIT intervals began with a 1:2 WRR and progressed to a 3:4 WRR in the final week of the intervention. This adjustment aligns with the rise in fitness and thus ensures that the target HR between 80% and 95% of MaxHR is consistently achieved.

The APEX program was developed to be an affordable and accessible exercise intervention fostering strong adherence among older adults. With the multimodal progressive HIIT format, APEX was uniquely designed to safely provide maximal physical and cognitive benefits to older adults, including those that may be deconditioned. The main goal of this study was to evaluate the feasibility of the novel APEX program in healthy older adults. Specifically, our first aim was to establish the safety of the intervention and gauge its adherence among healthy older adults. The second aim was to demonstrate the acute physiological effects of exercise in accordance with predefined parameters of the HIIT protocol. The third and final exploratory aim was to assess possible functional fitness and cognitive changes following the intervention.

## Methods

### Participants

Four community-dwelling and fully ambulatory older adults were recruited from community centers in a large metropolitan area (see Table 1 for Participant information). No participants had any medical issues that might create a safety issue for exercise, as determined by the PAR-Q+ health history questionnaire (Warburton, et al., 2011). No participants had a history of neurological conditions or reported using antidiabetic medications.

**Table 1:** Participant demographics.

| **ID** | **Age Range**<br>**(years)** | **Education**<br>**Level (years**<br>**of schooling)** | **Gender** | **Hours of**<br>**Physical**<br>**Activity per**<br>**Week** | **MaxHR**<br>**(bpm)** |
| --- | --- | --- | --- | --- | --- |
| E101 | 65-69 | 18 | Male | 5 | 142 |
| E102 | 75-79 | 14 | Female | 7 | 132 |
| E103 | 65-69 | 16 | Female | 5 | Not measured |
| E104 | 75-79 | 16 | Male | 6 | 157 |
| Mean | 71.3 ± 5.2 | 16 ± 1.6 | N/A | 5.3 ± 0.6 | 143.7 |

*Note. MaxHR - maximum heart rate.*

For three of the participants (E101, E102, and E104), VO_2_Max and MaxHR were measured via indirect calorimetry utilizing a treadmill ramp protocol (Pescatello et al., 2014). For the remaining participant (E103), VO_2_Max and MaxHR could not be measured due to scheduling conflicts. Prior to recruitment, the study was approved by the authors’ Institutional Review Boards. All participants provided written informed consent before participation. The study was registered on ClinicalTrials.gov (NCT06199661).

### Study design

This study utilized a pre-test post-test treatment design. Participants were assessed with a set of functional fitness and cognitive measures before and after the exercise program. Since the format of the computerized cognitive tasks was not familiar to the participants, we anticipated that there might be some improvements unrelated to any real cognitive gains between the first and second assessments due to learning the tasks. Thus, we utilized a multiple-baseline paradigm for cognitive measures and added an additional baseline session. The APEX program (described in detail below) was used as the exercise intervention, performed in an in-person group setting.

Overall, the sequence of study events was the following:

- Participant recruitment, screening, and consent;
- Baseline testing session (#1), cognitive measures only (a week before the Pre-exercise session);
- Pre-exercise session (#2), VO_2_Max and MaxHR assessment, functional fitness and cognitive measures (3-7 days before the start of the exercise program);
- APEX program, two in-person group classes per week for four weeks, eight classes in total;
- Post-exercise session (#3), functional fitness and cognitive measures, (0-2 days after the end of the exercise program).

### Physical exercise intervention

APEX is a multimodal progressive HIIT program aimed to safely elicit multifaceted improvements in functional fitness and cognition in older adults. The APEX program had three phases: warm-up, HIIT (repeated three times per class), and cooldown (detailed below in Table 2). The 10-minute warm-up included deep breathing meditation and dynamic joint mobility exercises (Table 2). Then, participants completed a 40-minute strength and balance workout in HIIT format with five different high-intensity (HI) exercises interspersed with five different postural stability recovery exercises. The HI intervals focused on enhancing cardiovascular fitness, muscle strength, and motor performance. The recovery intervals incorporated balance and mobility exercises. The HIIT intervals began with a 1:2 WRR and progressed to a 3:4 WRR in the final week. Specifically, the HI exercises lasted 30 seconds for the first three weeks and then increased to 45 seconds in the fourth week while the recovery exercises lasted 60 seconds throughout the entire intervention (Table 2). This adjustment in WRR aligned with the rise in fitness, ensuring that the target HR between 80% and 95% of MaxHR was consistently achieved. This HIIT phase of ten exercises was repeated three times in the 40 minutes (except for the first class, in which the specific movements associated with each exercise were introduced to the participants for the first time, which required additional time, and thus only two repetitions were accomplished). Each APEX class ended with a 10-minute cooldown consisting of standing and seated exercises similar to the warm-up (Table 2). Participant E103 engaged in two APEX classes remotely over Zoom, due to predetermined scheduling conflicts.

The exercises for APEX aimed to increase oxygen transport to larger muscles through repetitive, continuous movements, like hip steps, marches, and feet rocking. This ensured that the post-APEX HR was genuinely elevated due to increased oxygen transport, not artificially increased by a sympathetic nervous system response (Snarr, 2022). This guaranteed that participants’ VO_2_ and HR consistently adhered to HIIT protocols.

To monitor both the safety of participants and the extent of their exertion during the intervention (i.e., evaluate its acute physiological effects), participants’ HRs were measured. HRs were measured using Polar 810i HR monitors. In the rare instances in which automatic measurement was not possible, HRs were manually palpated from the radial artery over 15 seconds. The target HR range for high-intensity intervals was at or greater than 80% of measured MaxHR, and the target HR range for recovery intervals was below 80%. HRs were taken immediately after each high-intensity exercise, each recovery exercise, the warm-up phase, and the cooldown phase for every class.

**Table 2:**
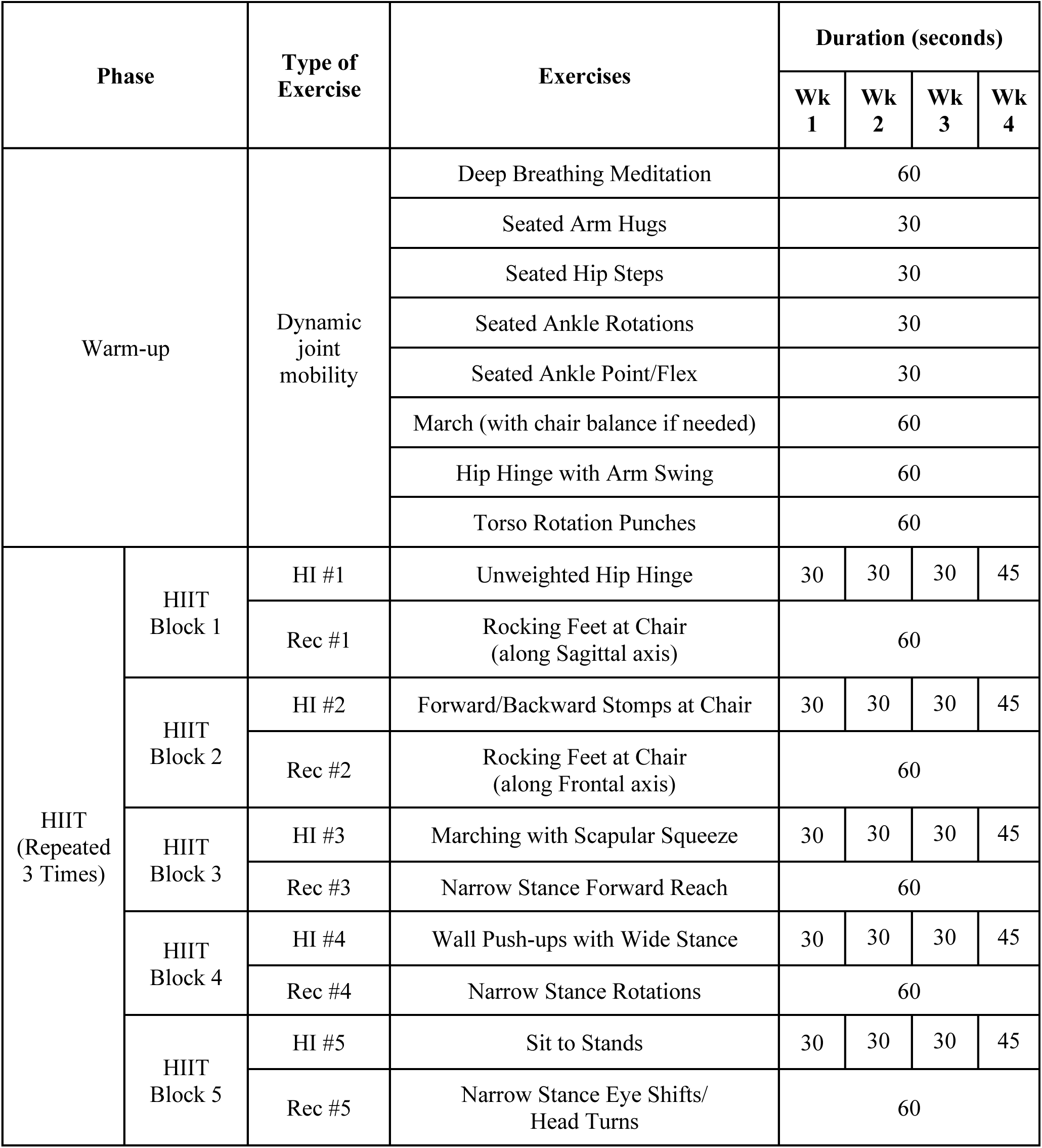
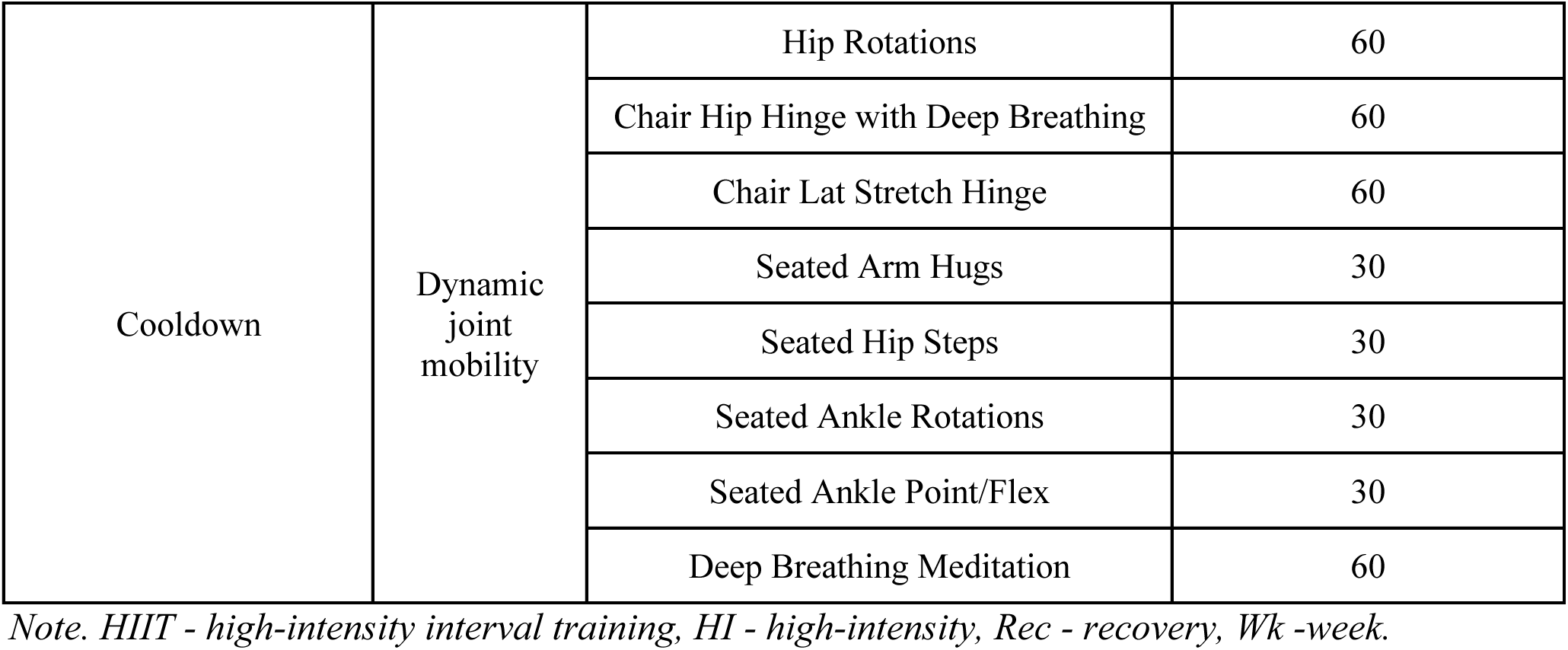
Detailed outline of the APEX training.

| Phase |  | Type of Exercise | Exercises | Duration (seconds) |  |  |  |
| --- | --- | --- | --- | --- | --- | --- | --- |
|  |  |  |  | Wk 1 | Wk 2 | Wk 3 | Wk 4 |
| Warm-up |  | Dynamic joint mobility | Deep Breathing Meditation | 60 |  |  |  |
|  |  |  | Seated Arm Hugs | 30 |  |  |  |
|  |  |  | Seated Hip Steps | 30 |  |  |  |
|  |  |  | Seated Ankle Rotations | 30 |  |  |  |
|  |  |  | Seated Ankle Point/Flex | 30 |  |  |  |
|  |  |  | March (with chair balance if needed) | 60 |  |  |  |
|  |  |  | Hip Hinge with Arm Swing | 60 |  |  |  |
|  |  |  | Torso Rotation Punches | 60 |  |  |  |
| HIIT<br>(Repeated 3 Times) | HIIT Block 1 | HI #1 | Unweighted Hip Hinge | 30 | 30 | 30 | 45 |
|  |  | Rec #1 | Rocking Feet at Chair<br>(along Sagittal axis) | 60 |  |  |  |
|  | HIIT Block 2 | HI #2 | Forward/Backward Stomps at Chair | 30 | 30 | 30 | 45 |
|  |  | Rec #2 | Rocking Feet at Chair<br>(along Frontal axis) | 60 |  |  |  |
|  | HIIT Block 3 | HI #3 | Marching with Scapular Squeeze | 30 | 30 | 30 | 45 |
|  |  | Rec #3 | Narrow Stance Forward Reach | 60 |  |  |  |
|  | HIIT Block 4 | HI #4 | Wall Push-ups with Wide Stance | 30 | 30 | 30 | 45 |
|  |  | Rec #4 | Narrow Stance Rotations | 60 |  |  |  |
|  | HIIT Block 5 | HI #5 | Sit to Stands | 30 | 30 | 30 | 45 |
|  |  | Rec #5 | Narrow Stance Eye Shifts/<br>Head Turns | 60 |  |  |  |

|  |  |  |  |
| --- | --- | --- | --- |
| Cooldown | Dynamic joint mobility | Hip Rotations | 60 |
|  |  | Chair Hip Hinge with Deep Breathing | 60 |
|  |  | Chair Lat Stretch Hinge | 60 |
|  |  | Seated Arm Hugs | 30 |
|  |  | Seated Hip Steps | 30 |
|  |  | Seated Ankle Rotations | 30 |
|  |  | Seated Ankle Point/Flex | 30 |
|  |  | Deep Breathing Meditation | 60 |

*Note. HIIT - high-intensity interval training, HI - high-intensity, Rec - recovery, Wk -week.*

### Assessment measures

*Functional fitness measures.* During the Session 2 (pre-exercise) and Session 3 (post-exercise) testing, participants underwent a 2-minute Step Test to assess aerobic endurance, 30 seconds of Chair Stands to quantify lower body strength, an 8-foot Timed Up-and-Go test to gauge agility and dynamic balance, and a Functional Reach Test.

#### Functional Reach Test

The Functional Reach Test (FRT) is an assessment of static balance and joint mobility (Duncan et al., 1990; Thompson et al., 2023). The FRT measures the distance (± 0.25 inches) of an individual’s forward reach with one arm while maintaining a standing position with feet flat on the floor at hip width (Duncan et al., 1990).

#### Timed Up-And-Go

The Timed Up-And-Go test (TUG) is an assessment of agility and dynamic balance and is often used to assess functional mobility, especially in older adults (Podsiadlo & Richardson, 1991).

The TUG test measures the number of seconds (± 0.01 second) for an individual to rise from a seated position, walk eight feet, turn around, and return to a seated position (Podsiadlo & Richardson, 1991).

#### 2-minute Step Test

The 2-minute Step Test is an assessment of functional mobility and aerobic endurance (Jones et al., 1999; Thompson et al., 2023). The 2-minute Step Test measures the number of right knee raises completed in two minutes while marching in place and raising each knee to a point midway between the patella (kneecap) and iliac crest (top hip bone).

#### 30-Second Chair Stand Test

The 30-Second Chair Stand Test (30CST) assesses lower body strength (Jones et al., 1999; Thompson et al., 2023). The 30CST measures the number of sit-to-stands completed in 30 seconds on a standard 17” seat-height chair with arms folded across the chest.

##### Cognitive measures

Eight cognitive tasks testing information processing, working memory, executive control, and attention were administered to the participants three times, twice before the intervention and once after (see Study Design section for more details). To familiarize participants with the interface for the cognitive tasks, the task with the easiest instructions was shown first (Visual Search Task). The remaining tasks were presented in a random order each time.

The cognitive tasks were created using the software PsyToolkit, which allows researchers to code their own computerized experiments and automatically record participants’ responses to each trial (Stoet, 2010; Stoet, 2017). The participants completed the tasks on a computer with an experimenter present to facilitate the instructions and training trials.

#### Information Processing

The Visual Search Task is an information processing test that requires participants to respond with a keypress if a target symbol is present in a collection of 5, 10, 15, or 20 symbols, but refrain from responding if no target symbol is present (Treisman & Gelade, 1980). The dependent measures were the participant’s accuracy per condition (5, 10, 15, or 20 items) and their reaction time on correct trials per condition.

#### Working Memory Capacity Tasks

The Forward Digit Span Task is a working memory task that requires participants to recall a sequence of digits that successively flash on-screen (Jones, 2015). In the first trial, two digits successively flash on the screen. If the participant correctly recalls the two-digit sequence, they are prompted with another random two-digit sequence. If they again correctly recall this new sequence, the next sequence is of length three, and so on. Participants responded by clicking digits from an on-screen number pad. The dependent measure was the participant’s digit span— the longest sequence length they could correctly recall two times.

Similarly, the Backward Digit Span Task is a working memory task that requires participants to report the reverse of a sequence of digits that successively flash on-screen (e.g., if the stimulus is “1-2-3”, the correct response would be “3-2-1”). The dependent measure was the participant’s backward digit span—the longest sequence length for which the participant could correctly report backward two times.

The Forward Spatial Span Task is a working memory task that requires participants to recall and click a sequence of random positions, designated by squares that “light up” on the screen (Corsi, 1972). In the first trial, two squares light up successively. The number of squares that light up in the sequence increases by one with each trial. The dependent measure was the participant’s spatial span, the longest sequence the participant could recall correctly.

The Backward Spatial Span Task requires participants to recall and select a sequence of random positions on the screen in backward order (Isaacs & Vargha-Khadem, 1989; Kessels et al., 2008). The dependent measure was the participant’s backward spatial span, the longest sequence the participant can recall correctly.

#### Executive Control and Attention Tests

The Go/No-Go Task is an inhibitory control task that requires participants to respond with a keypress when a “Go” is presented on the screen but refrain from responding when a “No Go” is presented (Verbruggen & Logan, 2008). In our task, the ratio of Go trials to No-Go trials was 4:1 and there was a short intertrial delay of 450 milliseconds, designed to maximize false alarms— incorrectly responding with a keypress for a No-Go trial (Young et al., 2018). The dependent measures were the participant’s hits—the proportion of correct responses on the Go trials—and their reaction times on hit trials.

The Flanker Task is a focused attention and interference control test that requires participants to respond with a left arrow keypress or right arrow keypress corresponding to the direction of the arrow in the center of five symbols (Eriksen & Eriksen, 1974). In congruent trials, all of the symbols point in the same direction (e.g., for “>>>>>”, the correct response is the right arrow key). In incongruent trials, the flanking arrows are pointing in the opposite direction of the central arrow (e.g., for “<<><<”, the correct response is the right arrow key). In neutral trials, the flanking symbols are dashes (e.g., for“-->--”, the correct answer is the right arrow key). The dependent measures were the participant’s accuracy by condition (congruent, incongruent, neutral) and their reaction times on correct trials per condition.

The Stroop Task is a focused attention and interference control test that requires participants to respond with the font color of a presented word (Stroop, 1935). The presented words are names of different colors: *‘yellow’*, *‘blue’*, *‘red’*, or *‘green’*. The task includes congruent trials, where the written word matches its font color (e.g., word *‘blue’* written in blue font), and incongruent trials, where they do not match (e.g., word *‘red’* written in blue font). In both instances, the participants must identify the font color (blue in both given examples) and press the key corresponding to the first letter of the font color name (e.g., the ‘b’ key for blue); this is inherently more difficult in incongruent trials. The dependent measures were the participant’s accuracy by condition (congruent or incongruent) and their reaction times on correct trials by condition.

### Statistical analysis

First, one-tailed one-sample t-tests were used to evaluate whether measured HRs following each high-intensity exercise were significantly lower than 80% of MaxHR across all participants with MaxHR data. A one-tailed test was used because the goal of the high-intensity exercises was to reach a HR equal to or greater than the predefined HR threshold. A lack of significance in this case indicates that the measured HR was greater than or not statistically different from the target threshold of 80% MaxHR. Second, one-tailed one-sample t-tests were conducted to ascertain whether recovery HRs were significantly lower than 80% of MaxHR. Third, linear mixed-effects models were used to evaluate whether a significant difference existed between HRs collected during high-intensity and recovery periods for each HIIT block, with interval type (high-intensity vs. recovery) used as a fixed factor and participant, session, and repetition number as random factors. An overall analysis across all blocks was also run with interval type as a fixed factor and participant, session, repetition number, and block as random factors. Last, we explored these patterns on an individual level to determine whether the training similarly impacted individual participants. Again, we used one-tailed one-sample t-tests to compare measured HRs following each high-intensity exercise and recovery exercise to 80% of MaxHR. We also ran paired two-sample t-tests to compare recovery and high-intensity HRs.

For all HR data analyses, data from the first APEX class were excluded since this class familiarized participants with the exercises. An alpha cutoff of 0.05 was used for all statistical tests. Finally, we explored changes in functional fitness and cognitive measures from pre-to post-exercise. Only descriptive statistics are provided for these comparisons, as statistical tests could not be performed due to the small sample size. All data analyses were performed using R Statistical Software (ver. 4.3.1; R Core Team, 2023) and figures were drawn in ggplot2 (ver. 3.4.3; Wickham, 2016). The linear mixed-effects model analyses were performed with the lme4 package (ver. 1.1-34; Bates et al., 2015).

## Results

### Safety and feasibility of the APEX program

No adverse events were observed for any participant during their participation in the APEX program. Adherence to the program was high; three participants attended all eight APEX classes, and one participant attended seven out of eight APEX classes due to a scheduling conflict that was known at the time of enrollment.

### Acute physiological effects

For the high-intensity exercises, we evaluated whether the average HRs were greater than or equal to the threshold 80% of a participant’s MaxHR. Based on one-tailed one-sample t-tests, none of the mean HRs for the high-intensity exercises were found to be significantly lower than 80% of MaxHR across all participants with MaxHR data (E101, E102, E104) (as evidenced by p-values > 0.05, see Table 3). Moreover, three high-intensity exercises—Forward/Backward Stomps at Chair, Marching with Scapular Squeeze, and Sit to Stands—elicited HRs that significantly exceeded the 80% threshold on average (Table 3).

**Table 3:**
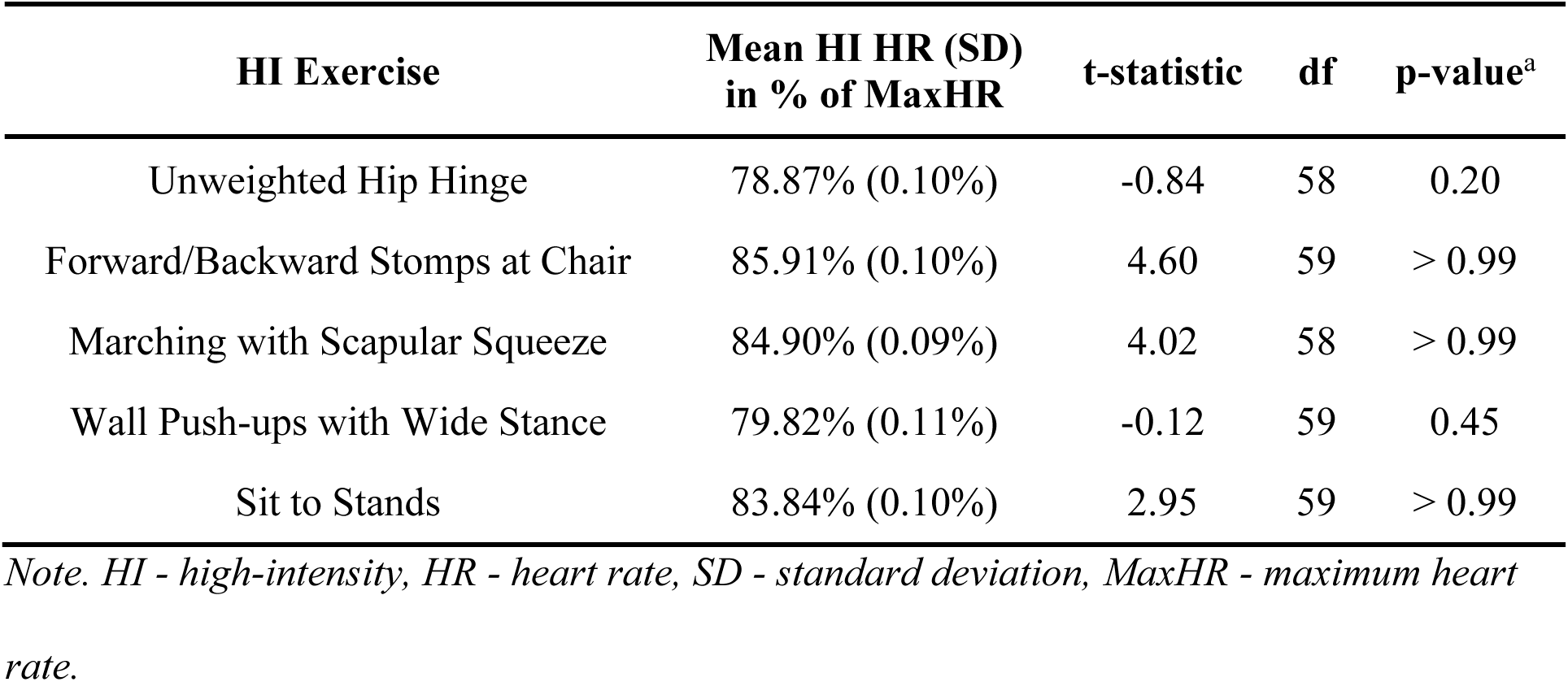
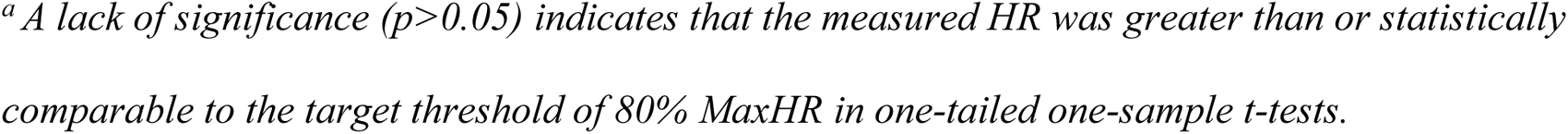
Descriptive statistics for measured heart rates after high-intensity exercises and results of one-tailed one-sample t-tests comparing them to threshold heart rate of 80% of maximum heart rate.

| HI Exercise | Mean HI HR (SD)<br>in % of MaxHR | t-statistic | df | p-value<sup>a</sup> |
| --- | --- | --- | --- | --- |
| Unweighted Hip Hinge | 78.87% (0.10%) | -0.84 | 58 | 0.20 |
| Forward/Backward Stomps at Chair | 85.91% (0.10%) | 4.60 | 59 | > 0.99 |
| Marching with Scapular Squeeze | 84.90% (0.09%) | 4.02 | 58 | > 0.99 |
| Wall Push-ups with Wide Stance | 79.82% (0.11%) | -0.12 | 59 | 0.45 |
| Sit to Stands | 83.84% (0.10%) | 2.95 | 59 | > 0.99 |

*Note. HI - high-intensity, HR - heart rate, SD - standard deviation, MaxHR - maximum heart rate.*
<sup>a</sup> A lack of significance ( $p > 0.05$ ) indicates that the measured HR was greater than or statistically comparable to the target threshold of 80% MaxHR in one-tailed one-sample t-tests.

For the recovery exercises, we determined whether the average HR was below 80% of MaxHR. According to one-tailed one-sample t-tests, four out of the five recovery exercises elicited mean HRs that were significantly lower than 80% of MaxHR across all participants with MaxHR data (Table 4). Only the Narrow Stance Forward Reach exercise was non-significantly lower than the indicated threshold.

**Table 4:**
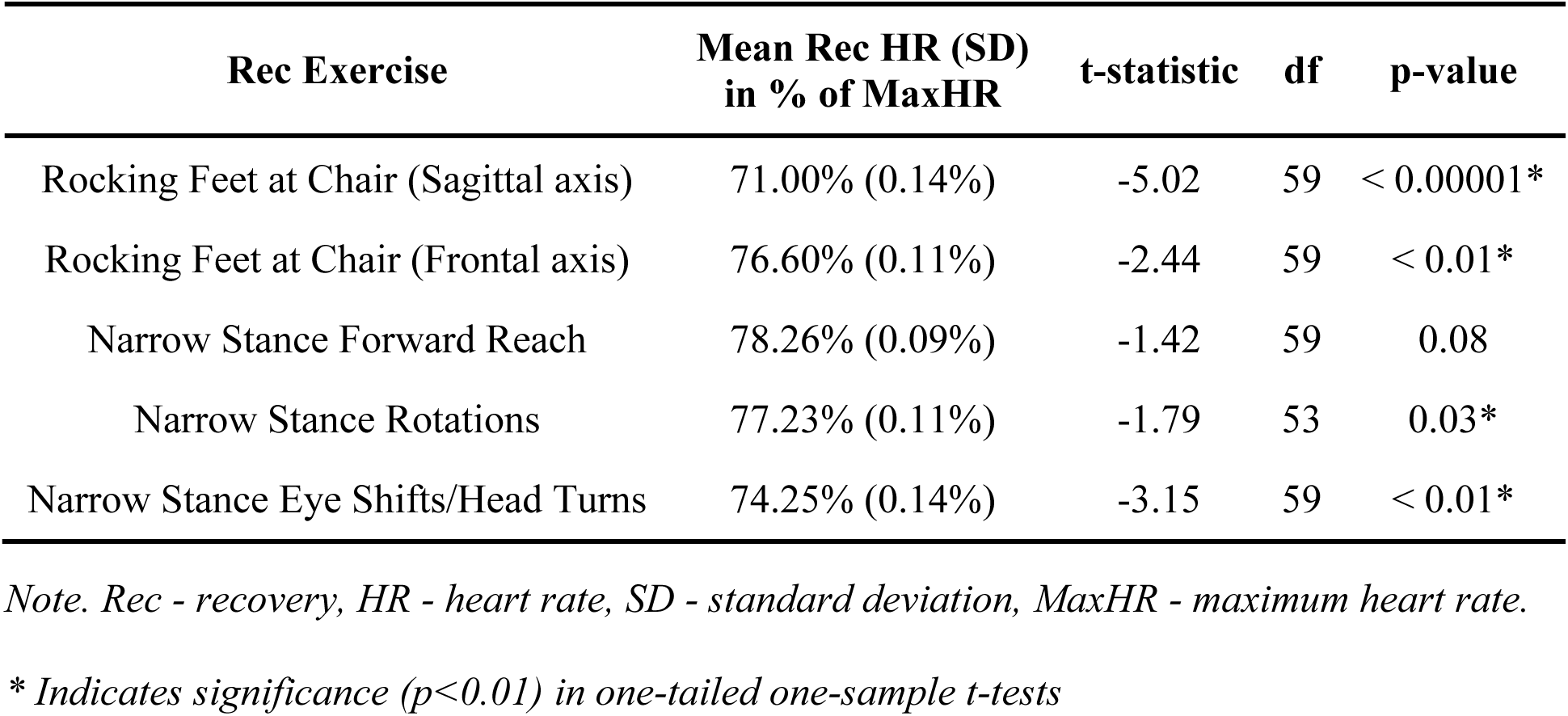
Descriptive statistics for measured heart rates after recovery exercises and results of one-tailed one-sample t-tests comparing them to threshold heart rate of 80% of maximum heart rate.

Next, using the linear mixed-effects model, we showed that across all participants and APEX classes, there was a statistically significant difference between the HRs of high-intensity and recovery periods for each HIIT block and across all HIIT blocks (Table 5). HIIT Block 4 (HI exercise: Wall Push-Ups with Wide Stance, recovery exercise: Narrow Stance Rotations) had the smallest difference between HI and recovery HRs at 4.99 beats per minute. Overall, there was an average difference of 11.41 beats per minute between the high-intensity and recovery HRs.

**Table 5:** Results of linear mixed-effects models comparing heart rates between high-intensity and recovery periods for each HIIT block and across all blocks.

| Exercise | Fixed effects | Estimates | SE | t-statistic | df | p-value |
| --- | --- | --- | --- | --- | --- | --- |
| HIIT Block 1 | Intercept | 99.34 | 5.41 | 18.37 | 4.56 | < 0.001* |
|  | Interval (HI vs Rec) | 12.35 | 2.09 | 5.92 | 146.12 | < 0.001* |
| HIIT Block 2 | Intercept | 107.70 | 5.64 | 19.12 | 3.95 | < 0.001* |
|  | Interval (HI vs Rec) | 13.75 | 1.82 | 7.58 | 148.87 | < 0.001* |
| HIIT Block 3 | Intercept | 110.42 | 5.72 | 19.31 | 4.20 | < 0.001* |
|  | Interval (HI vs Rec) | 10.75 | 1.67 | 6.45 | 147.95 | < 0.001* |
| HIIT Block 4 | Intercept | 108.14 | 7.03 | 15.38 | 3.56 | < 0.001* |
|  | Interval (HI vs Rec) | 4.99 | 1.94 | 2.58 | 143.49 | < 0.001* |
| HIIT Block 5 | Intercept | 104.13 | 6.31 | 16.51 | 4.03 | < 0.001* |
|  | Interval (HI vs Rec) | 14.90 | 2.17 | 6.88 | 150.81 | < 0.001* |
| Overall | Intercept | 105.94 | 6.25 | 16.94 | 4.73 | < 0.001* |
|  | Interval (HI vs Rec) | 11.41 | 0.87 | 13.19 | 781.09 | < 0.001* |

*Note. HIIT - high-intensity interval program, HI - high-intensity, Rec - recovery, SE - standard error.*
*\* Indicates significance ( $p < 0.001$ )*

Finally, we analyzed HR data on an individual level. For two of the three participants with MaxHR data, their individual mean HRs consistently met or exceeded the threshold HRs in all of the high-intensity exercises in one-tailed one-sample t-tests. Only one participant’s (E101) mean HR was significantly lower than the threshold HR for the Unweighted Hip Hinge and Marching with Scapular Squeeze high-intensity exercise; no other participants’ mean HRs were significantly lower than their threshold HR in any of the APEX high-intensity exercises (Figure 1). Next, using paired two-sample t-tests, individual recovery HRs were found to be significantly lower than high-intensity HRs across all participants and HIIT blocks, except for E101 in HIIT block #4 (Wall Push-ups with Wide Stance and Narrow Stance Rotations) (Figure 1). For corresponding values and statistical values, see Appendix, Tables A1 and A2.

**Figure 1:**
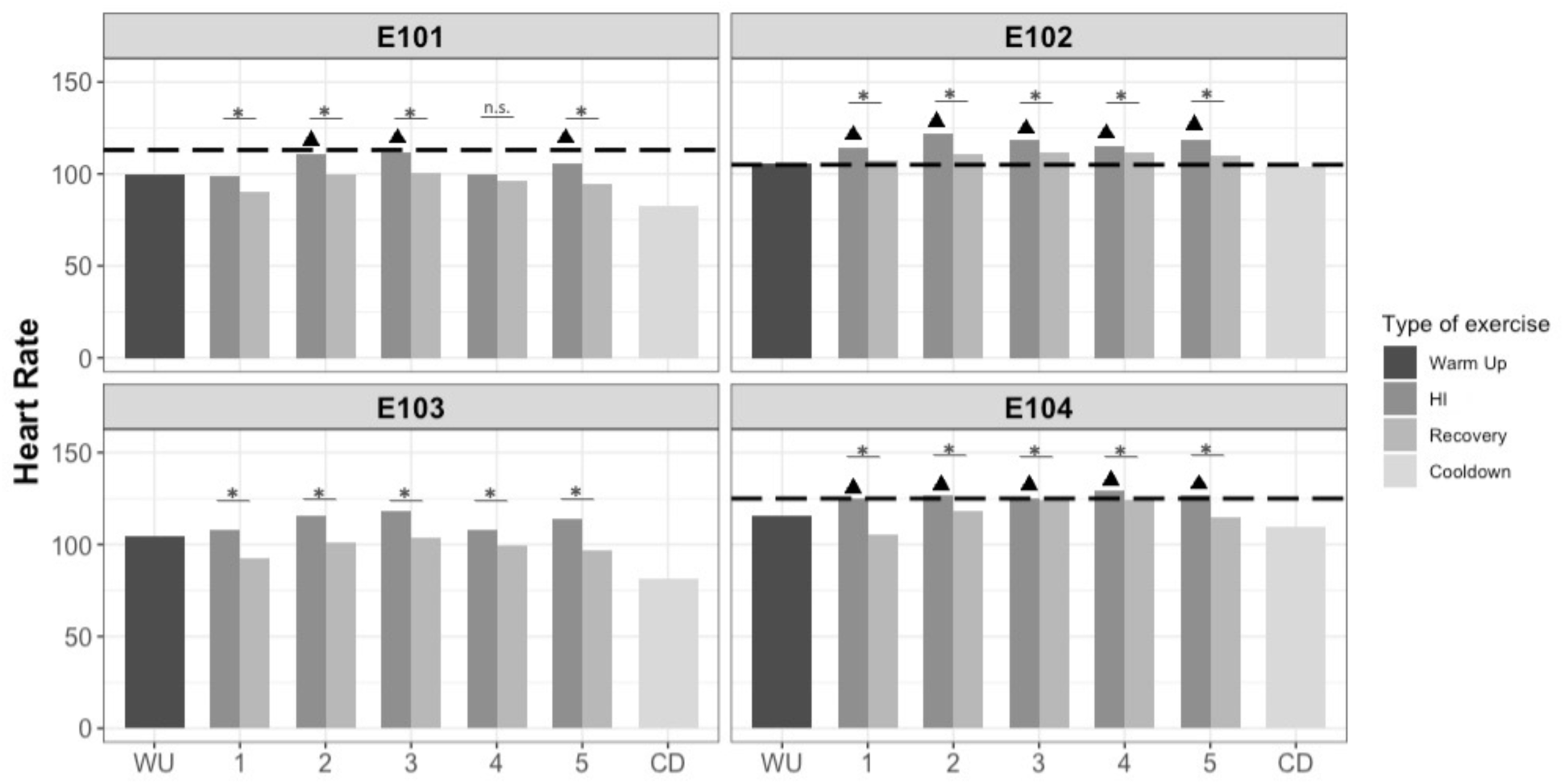
Individual mean heart rates during APEX training. Heart rates during warm up (WU), each HIIT block’s high-intensity (HI) and recovery exercise, and cooldown (CD). Dotted lines indicate the individual threshold heart rate (HR) for the participant, calculated as 80% of maximum HR (MaxHR) for E101, E102, and E104. No MaxHR data was available for E103. The target HR range for HI exercise is at or above the threshold; the target recovery HR range is below the threshold. Triangles (▲) represent HI HRs statistically comparable to or greater than the threshold HRs. * Indicates significant (p<0.05) difference between HI and recovery exercise within a given HIIT block, as determined by paired two-sample t-tests (see Appendix, Tables A1 and A2 for actual values and test statistics).

### Functional fitness measures

Due to the small sample size, we are only able to provide descriptive statistics of changes from pre-to post-intervention on these measures. On the functional fitness tests, participants improved on the 2-Minute Step Test, Chair Stand Test, Timed Up-and-Go Test, and Functional Reach Test with the right arm, on average. The Functional Reach Test with the left arm showed a small decrease of 0.08 inches, on average (Table 6).

**Table 6:** Descriptive statistics of changes in functional fitness measures.

| **Test** | **Pre-APEX Mean**<br>**(SD)** | **Post-APEX Mean**<br>**(SD)** | **Mean Difference**<br>**(SD)** |
| --- | --- | --- | --- |
| FRT: Right arm | 13.42 in (2.01) | 14.50 in (1.81) | +0.5 in<sup>†</sup> (1.5) |
| FRT: Left arm | 15.08 in (2.90) | 15.19 in (1.91) | -0.08 in (1.94) |
| 2-Minute Step Test | 103.67 steps (2.08) | 119.25 steps (8.46) | +12.67 steps<sup>†</sup> (8.33) |
| Mean TUG | 7.13 sec (1.44) | 6.06 sec (0.83) | -1.07 sec<sup>†</sup> (1.27) |
| Chair Stand Test | 14.00 reps (1.00) | 17.00 reps (2.45) | +2.00<sup>†</sup> reps (1.00) |

*Note. FRT - Functional Reach Test, TUG - Timed Up-and-Go Test, SD - standard deviation.*
*<sup>†</sup> Indicates improvement in functional fitness between sessions. Due to the small sample size, no statistical tests were performed.*

### Cognitive measures

The accuracies for the Visual Search, Flanker, and Stroop Tasks were above 85% for all participants and all sessions. Since all participants were already at ceiling for these tasks (between 92% and 100%) in Session 1, we focused our analysis on changes in reaction times.

Participants’ Session 2 (pre-exercise) reaction times were faster than those of Session 1 (baseline) for all information processing, executive control, and attention tasks, and Session 2 spans were greater than Session 1’s for all working memory tasks except Forward Spatial Span. The improvements observed between Sessions 1 and 2 were likely due to learning effects as participants were familiarized with the tasks. This is in contrast to the differences in performance between Session 2 (pre-exercise) and Session 3 (post-exercise), which were more likely due to the effects of APEX, given that the participants were already familiar with the tasks. Improvements in reaction times or spans between Sessions 2 and 3 were observed in the Visual Search Task, Forward Digit Span Task, Forward Spatial Span Task, Go/No Go Task, and Flanker Task (incongruent condition) (Table 7).

**Table 7:** Descriptive statistics of changes in performance on cognitive tasks.

| **Cognitive Domain Tested** | **Tasks** | **S1 Mean Span or RT (SD)** | **S2 Mean Span or RT (SD)** | **S3 Mean Span or RT (SD)** | **S2 - S1 Avg Difference** | **S3 - S2 Avg Difference** |
| --- | --- | --- | --- | --- | --- | --- |
| **Info Processing**<br>(in ms) | Visual Search Task | 1836.43 ms<br>(615.52) | 1618.00 ms<br>(470.17) | 1576.41 ms<br>(630.19) | -218.43 ms<sup>†</sup> | -41.59 ms<sup>†</sup> |
| **Working Memory<sup>a</sup>**<br>(span size) | Forward DS | 4.75 (0.50) | 5.00 (0.82) | 5.75 (1.26) | +0.25<sup>†</sup> | +0.75<sup>†</sup> |
|  | Backward DS | 3.50 (0.58) | 4.25 (1.50) | 3.75 (0.50) | +0.75<sup>†</sup> | -0.50 |
|  | Forward SS | 4.50 (0.58) | 4.00 (0.82) | 4.75 (1.50) | -0.50 | +0.75<sup>†</sup> |
|  | Backward SS | 3.50 (0.58) | 4.75 (0.50) | 3.75 (0.50) | +1.25<sup>†</sup> | -1.00 |
| **Executive control & Attention<sup>b</sup>**<br>(in ms) | Go/No-Go Task | 577.89 ms<br>(192.13) | 507.48 ms<br>(141.50) | 478.27 ms<br>(157.14) | -70.41 ms<sup>†</sup> | -29.20 ms<sup>†</sup> |
|  | Flanker Task (Congruent) | 799.52 ms<br>(160.51) | 711.90 ms<br>(183.94) | 702.96 ms<br>(231.57) | -87.61 ms<sup>†</sup> | -8.94 ms<sup>†</sup> |
|  | Flanker Task (Incongruent) | 916.62 ms<br>(275.69) | 835.85 ms<br>(140.87) | 771.47 ms<br>(185.32) | -80.77 ms<sup>†</sup> | -64.38 ms<sup>†</sup> |
|  | Flanker Task (Neutral) | 785.77 ms<br>(236.11) | 678.18 ms<br>(122.88) | 693.44 ms<br>(153.76) | -107.59 ms<sup>†</sup> | +15.26 ms |
|  | Stroop Task (Congruent) | 1308.37 ms<br>(116.60) | 1167.37 ms<br>(317.81) | 1179.73 ms<br>(349.19) | -140.99 ms<sup>†</sup> | +12.36 ms |
|  | Stroop Task (Incongruent) | 1421.36 ms<br>(183.14) | 1252.57 ms<br>(313.81) | 1257.96 ms<br>(344.92) | -168.79 ms<sup>†</sup> | +5.40 ms |

*Note. S1 - session 1, S2 - session 2, S3 - session 3, RT - reaction time, SD - standard deviation,*
*DS - Digit Span, SS - Spatial Span*
*† Indicates improvement in span or reaction time between sessions. Due to the small sample size, no statistical tests were performed.*
*<sup>a</sup> Larger values indicate greater working memory capacity*
*<sup>b</sup> Shorter reaction times indicate faster responses and likely reflect more efficient processing capabilities*

## Discussion

This pilot study affirmed the APEX training’s safety for healthy older adults, with no adverse events. The routine also proved feasible for this population, resulting in high adherence and no attrition. One participant successfully attended two APEX classes over Zoom, further demonstrating its adaptability to diverse settings, including remote delivery.

HR measurements were used in this study to monitor whether APEX was eliciting the desired physiological effects during the high-intensity and recovery periods. Target HRs for high-intensity exercises were set at or above a threshold of 80% of MaxHR, and target HRs for recovery exercises were below that threshold. For all of APEX’s high-intensity exercises, the group mean HRs either surpassed or were statistically comparable to the threshold. These results signify that all of the high-intensity exercises evoked increased HRs to the desired level (at or above 80% MaxHR). Similarly, all recovery exercises elicited mean HRs in the desired recovery HR range (below 80% of MaxHR), and four out of the five recovery exercises induced HRs significantly below that threshold. Furthermore, all HIIT blocks demonstrated statistically significant differences in HRs between high-intensity and recovery periods.

Some high-intensity exercises were more effective at increasing heart rate than others. Specifically, the Unweighted Hip Hinge and the Wall Push-ups with Wide Stance exercises elicited a lower heart rate response than the other three exercises. These two exercises did not involve the lower body to the same extent as the others that involved marching, stomping, and/or repetitive flexion/extension motions. Less lower body engagement therefore decreased oxygen demand and heart rate response. Overall, the mean HRs of our study were consistent with similar HIIT studies, which found that high-intensity exercises elicited HRs exceeding 80% of MaxHR during high-intensity intervals for older adults (Elboim-Gabyzon et al., 2021; Gibala et al., 2018; Snarr, 2022).

The individual HR data reveal that two out of the three participants with MaxHR data had HRs that consistently met or exceeded the threshold of 80% of MaxHR for the high-intensity periods, with HRs significantly decreasing in the subsequent recovery periods. Even for the singular participant who did not consistently attain the targeted HRs (E101), a significant difference between high-intensity and recovery HRs was observed in four out of the five HIIT blocks. It is possible E101 did not exert himself adequately, as he subjectively reported lower exertion than other participants throughout the intervention.

On functional fitness tests, improvements following the APEX training were observed in the 2-Minute Step Test (assessment of aerobic endurance), Chair Stand Test (assessment of lower body strength), Timed Up-and-Go Test (assessment of agility and gait), and the Functional Reach Test (assessment of static balance and joint mobility) for the right arm, although no inferential statistics could be performed. These results are consistent with prior HIIT research, which overwhelmingly note significant exercise-induced improvements in aerobic endurance and lower body strength (Gibala et al., 2018; Marriott et al., 2021; Stern et al., 2023). Some HIIT studies have also found improvements in agility, gait, and dynamic balance in older adults (Elboim-Gabyzon et al., 2021; Gibala et al., 2018; Marriott et al., 2021, Stern et al., 2023). Previous studies that incorporated multimodal progressive training into HIIT routines showed similar findings, primarily supporting benefits for strength and endurance (Briggs et al., 2018; Buckley, et al., 2015; Lichtenstein et al., 2020; Sharp et al., 2022). Both HIIT and multimodal progressive literature have some limited support for HIIT improving static balance and joint mobility in older adults (Marriott et al., 2021; Stern et al., 2023). APEX may yield more pronounced functional fitness effects by targeting a greater variety of functional fitness domains and expanding the intervention duration in future studies.

To explore the possible cognitive effects of APEX, participants completed cognitive tasks. After the exercise program, the participants demonstrated faster reaction times in the information processing task and some executive control and attention tasks, in addition to modest improvements in some working memory tasks. Although we were unable to test for statistical significance, the observed exercise-induced improvements in executive control and attention strongly align with those found in previous exercise studies involving healthy individuals and different clinical populations (Ai et al., 2021; Cumming et al., 2012; Kluding et al., 2011). We hypothesize that, with a longer duration of training and a larger cohort, these trends would reach statistical and clinical significance, especially considering that other studies have demonstrated exercise-induced cognitive benefits with interventions lasting eight weeks or more (Vanderbeken & Kerckhofs, 2017). Future studies could examine a longer intervention to test for long-term adverse effects and explore the physical and cognitive effects of chronic APEX exercise.

Although the APEX intervention already shows great promise, it could be made more effective with the following modifications that should be explored in future research. Firstly, training participants in proper movement and muscle engagement could enhance functional fitness and ensure more consistent attainment of target heart rates. Additionally, providing visual or auditory cues, either through metronome timing, guiding proper form and repetition targets, or continuous real-time monitoring of HRs, may better motivate participants to exert appropriate effort and perform exercises correctly, thus helping them reach HR intensity thresholds (Lee et al., 2022; Schaefer, 2014). Furthermore, training participants to accurately report their Rating of Perceived Exertion (RPE), a validated self-reported measure of exercise intensity based on the participant’s perception of physical exertion, could serve as an additional tool to monitor exercise intensity (Morishita et al., 2018). With sufficient training, participants could consistently provide accurate RPEs, and this measure may allow for more accurate insight into participants’ physical exertion levels throughout the session, as it is strongly correlated with objective measures like HR.

Given the participants’ comfortable tolerance of the exercises in this pilot study, subsequent versions of APEX may consider extending the duration of the high-intensity workouts to increase exertion levels and potentially enhance physical and cognitive benefits. In other HIIT studies, longer interval durations ranging from 1 to 4 minutes are most prevalent (Marriott et al., 2021). Still, it is advisable to retain the progressive model employed in the APEX training, in which the duration increases week to week, in future studies to ensure it remains accessible across different ability levels.

In this pilot study, the participants were healthy and active older adults, a demographic that might experience only modest gains in cognition compared to sedentary and clinical populations through such an intervention. Since APEX is designed to accommodate varying levels of functional fitness, it holds promise for individuals across various clinical populations. To attain a more comprehensive understanding of the physical and cognitive effects of APEX, it would be advantageous to test the proposed intervention with a larger sample size, including participants from diverse clinical populations.

In particular, there is a compelling rationale to explore the benefits of APEX for post-stroke patients (Crozier et al., 2018; Gjellesvik et al., 2021; Hugues et al., 2021). It has been suggested that HIIT routines like APEX could provide neurocognitive benefits for post-stroke individuals, promoting recovery (Crozier et al. 2018; Hugues et al., 2021). This is because HIIT has demonstrated comparable safety and feasibility for clinical populations with greater time efficiency compared to MICT, encouraging adherence and making it an ideal exercise intervention for clinical populations (Anjos et al., 2022; Alzar-Teruel et al., 2022; Crozier et al., 2018; Gibala et al., 2018; Gjellesvik et al., 2021; Marriott et al., 2021). The APEX intervention’s accessibility and demonstrated safety and feasibility encourage its testing in the post-stroke population. Furthermore, since about one-third of stroke patients have aphasia, it would be informative to evaluate if APEX affects language recovery in people with aphasia with a language assessment. The effects of exercise on language are not well understood, and it is an area that requires more research.

Other areas of interest for future research with the APEX training are mood, mental health, and sleep. Older adults and clinical populations are at an elevated risk of depression and sleep disorders (Casagrande et al., 2022; Centers for Disease Control and Prevention, 2022; Liu et al., 2023; Uomoto, 2023). In addition to its numerous physical and cognitive benefits, physical activity has been shown to contribute to improved mood, mental health, and sleep in cognitively healthy and clinical older adults (Alfini et al., 2020; Vanderlinden et al., 2020; Yao et al., 2021). Therefore, an investigation into APEX’s effects on mood, mental health, and sleep in older adults and clinical populations is warranted.

## Conclusion

This pilot study demonstrated that the APEX program is safe and feasible in healthy older individuals, eliciting the appropriate physiological response with HRs aligning with existing HIIT guidelines. APEX offers a promising and accessible alternative to conventional cardiovascular exercise modalities for older adults, potentially providing additional benefits in various functional fitness, including aerobic endurance, lower body strength, agility, gait, dynamic, static balance, and joint mobility. Notably, positive trends in the cognitive domains of information processing, working memory, executive control, and attention were also observed. These results encourage future testing of an extended APEX program in older adults and different clinical populations with physical and cognitive impairments.

## Data Availability

All data produced in the present study are available upon reasonable request to the authors.

## Acknowledgments

We would like to acknowledge Dr. Erica Pitsch for her valuable input on the design of the APEX program. We would also like to thank the undergraduate students assisting with data collection. As always, we are deeply grateful to all the participants for their time.

