## Appendix for "Pilot study of a high-intensity interval training program in older adults: Safety, feasibility, functional fitness and cognitive effects"

***Table A1: Individual mean heart rates after high-intensity exercises and results of one-tailed one-sample t-tests comparing them to threshold heart rate.***

| ID | Threshold HR[a] (bpm) | HI exercise # | Mean HR (bpm) | t-statistic | df | p-value[b] |
|---|---|---|---|---|---|---|
| | | 1 | 99.3 | -7.57 | 19 | < 0.0001* |
| | | 2 | 111.00 | -0.84 | 20 | 0.21 |
| E101 | 113 | 3 | 112.10 | -0.37 | 20 | 0.36 |
| | | 4 | 99.76 | -4.61 | 20 | < 0.0001* |
| | | 5 | 105.86 | -3.47 | 20 | < 0.01* |
| | | 1 | 114.62 | 6.09 | 20 | > 0.99 |
| | | 2 | 121.62 | 12.13 | 20 | > 0.99 |
| E102 | 105 | 3 | 118.48 | 9.52 | 20 | > 0.99 |
| | | 4 | 115.14 | 8.43 | 20 | > 0.99 |
| | | 5 | 118.81 | 10.39 | 20 | > 0.99 |
| | | 1 | 107.86 | | | |
| | | 2 | 115.38 | | | |
| E103 | N/A[c] | 3 | 118.10 | N/A[c] | N/A[c] | N/A[c] |
| | | 4 | 107.57 | | | |
| | | 5 | 113.86 | | | |
| | | 1 | 124.83 | -0.04 | 17 | 0.48 |
| | | 2 | 137.67 | 3.09 | 17 | > 0.99 |
| E104 | 125 | 3 | 135.41 | 2.41 | 16 | > 0.99 |
| | | 4 | 129.28 | 0.97 | 17 | 0.83 |
| | | 5 | 137.33 | 3.21 | 17 | > 0.99 |

*Note. HR - heart rate, HI - high-intensity.*

** Indicates significance ($p<0.01$) in one-tailed one-sample t-tests*

[a] *Threshold heart rates are calculated as 80% of the individual's maximum heart rate, after each high-intensity exercise in APEX.*

[b] *A lack of significance ($p>0.05$) indicates that the measured HR was greater than or statistically comparable to the target threshold of 80% MaxHR.*

[c] *Threshold HR and descriptive statistics were not calculated for E103 as no MaxHR data was available for this participant.*

***Table A2: Individual differences in heart rate between high-intensity and recovery periods and results of paired two-sample t-tests comparing them.***

| ID | HIIT Block # | Mean difference between HI and Rec HR (bpm) | t-statistic | df | p-value |
|---|---|---|---|---|---|
| E101 | 1 | 7.95 | 6.22 | 19 | < 0.0001* |
| | 2 | 11.29 | 7.66 | 20 | < 0.0001* |
| | 3 | 11.43 | 4.95 | 20 | < 0.0001* |
| | 4 | 3.63 | 1.88 | 18 | 0.08 |
| | 5 | 11.29 | 8.87 | 20 | < 0.0001* |
| E102 | 1 | 6.95 | 5.19 | 20 | < 0.0001* |
| | 2 | 10.90 | 5.91 | 20 | < 0.0001* |
| | 3 | 6.43 | 3.62 | 20 | < 0.01* |
| | 4 | 3.21 | 4.07 | 18 | < 0.01* |
| | 5 | 9.14 | 8.87 | 20 | < 0.0001* |
| E103 | 1 | 16.32 | 10.01 | 18 | 0.0001* |
| | 2 | 14.52 | 8.50 | 20 | < 0.0001* |
| | 3 | 14.05 | 5.50 | 20 | < 0.0001* |
| | 4 | 8.37 | 2.79 | 18 | 0.01* |
| | 5 | 17.38 | 11.40 | 20 | < 0.0001* |
| E104 | 1 | 19.17 | 3.59 | 17 | < 0.01* |
| | 2 | 19.06 | 17.74 | 17 | < 0.0001* |
| | 3 | 12.71 | 9.63 | 16 | < 0.0001* |
| | 4 | 4.50 | 5.73 | 15 | < 0.0001* |
| | 5 | 22.94 | 3.79 | 17 | < 0.01* |

*Note. HIIT - high-intensity interval training, HI - high-intensity, Rec - recovery HR - heart rate.*

** Indicates significance ($p<0.01$) in paired two-sample t-tests*
